# Understanding Personal Protective Equipment Use Among Companion Animal Veterinary Staff in the Context of Zoonoses: A Qualitative Study

**DOI:** 10.64898/2026.08.24.26361178

**Authors:** Claudia Grace Cohen, Charlotte Robin, John S P Tulloch

## Abstract

**Introduction:** Veterinary Professionals are at risk of contracting zoonoses, including potential emerging infections. Personal Protective Equipment (PPE) could reduce infection transmission risk, but use of it in the profession is low. Understanding Veterinary Professionals’ experiences with PPE could help identify facilitators and barriers, therefore informing a future strategy to improve usage.

**Methods:** Two focus group discussion with veterinary nurses and two with veterinarians took place at a tertiary small animal teaching hospital. With an interpretivist epistemology, transcriptions were inductively coded and analysed using thematic analysis.

**Results:** Veterinary Professionals frequently reported underusing PPE, despite significant zoonotic risk. Friction in interdisciplinary relationships negatively impacted veterinary professionals’ experiences with PPE: differing views on risk, policy and PPE, contrasting perceptions of each other, and challenges with communication impacted PPE decision-making. Barriers included an absence of initial recognition of risk, a staff culture of prioritising patient health over risk to self, friction in response to others’ PPE use or lack thereof, an overly complex policy, and a lack of cultural and institutional reaction to occupationally contracted zoonoses. Facilitators included effective inter and intradisciplinary communication and previous personal experience with serious zoonoses.

**Conclusion:** Veterinary Professionals’ experiences with PPE, are shaped by social and professional factors, such as interdisciplinary friction, perceptions of self and others, of risk and policy. Recommendations are for the findings to be used locally to embed PPE use into everyday practice. At a national level, the United Kingdom Health Security Agency (UKHSA) should use the findings to evaluate current policies regarding PPE and zoonoses in veterinary practice and produce a simplified national guidance, in collaboration with veterinary professionals with lived experience. Further research into General Practice Veterinary Professionals’ experiences and the interdisciplinary social dynamics is recommended.

**Impacts:**

- Veterinarians and veterinary nurses often do not use adequate PPE when handling animals, despite the high risk of contracting zoonoses from them. Their experiences with PPE in the context of zoonoses have not yet been explored to identify key barriers and facilitators thus far.
- Veterinarians’ and veterinary nurses’ experiences with PPE are shaped personally, socially and culturally throughout their career.
- Interdisciplinary friction, overly complex policy, lack of risk recognition and patient prioritisation over self both personally and institutionally are barriers to PPE use. Good inter and intradisciplinary communication and personal experience with serious zoonoses are facilitators to PPE use.

## Introduction

Veterinary professionals (VPs) face elevated risks of zoonotic disease exposure due to close contact with sick animals (Baker & Gray, 2009; Burling, 2018; United Kingdom Health Security Agency, 2014; Weese, Peregrine, & Armstrong, 2002a, 2002b), with one study reporting a 44.7% prevalence of suspected infections, primarily dermatophytosis and campylobacteriosis (Robin, Bettridge, & McMaster, 2017). These cases often serve as early indicators of emerging pathogens. Reducing transmission is crucial for occupational health and preventing wider spread.

Infection Prevention and Control (IPC) measures, often referred to as biosecurity in a veterinary setting, such as hand hygiene and appropriate use of personal protective equipment (PPE), can reduce zoonotic transmission between animals and VPs (Burling, 2018) (Australian Veterinary Association, 2025; Rabinowitz & Conti, 2013; Scheftel et al., 2010). However, previous research has identified low PPE use in VPs (Dowd, Taylor, Toribio, Hooker, & Dhand, 2013; Robin et al., 2017). In the United Kingdom (UK) the Health and Safety Executive (HSE) regulates workplace health and safety. While veterinary practices must align with professional standards set by the Royal College of Veterinary Surgeons (RCVS), individual veterinary businesses retain responsibility for designing and implementing their own specific infection control policies. While the RCVS accredited practice scheme mandates the presence of PPE policies, only 69% of practices in the UK are accredited to this scheme (Royal College of Veterinary Surgeons). The RCVS provides no industry specific biosecurity guidance, relying instead on external resources (American Animal Hospital Association, 2020; CVS; Davies Veterinary Specialists, 2020), and the HSE zoonoses guidance focuses on agriculture and laboratory work only, leaving companion animal practice without specific direction (Health and Safety Executive, 1997), except for specific guidance for veterinary practices on control measures for Brucella Canis (Animal and Plant Health Agency, 2025a).

Knowledge and availability of practice IPC guidelines may affect PPE use (Robin et al., 2017), however VPs may be unaware of guidelines and some workplaces lack IPC policies (Baiyasi et al.; Chakraborty, Fama, & Sander, 2024; Dowd et al., 2013; Lipton, Hopkins, Koehler, & DiGiacomo, 2008; Venkat, Yaglom, & Adams, 2019; Wright, Jung, Holman, Marano, & McQuiston, 2008). There is no clear consensus on how years in practice influences likelihood of contracting zoonoses or PPE use. One study found veterinary surgeons practicing for over 20 years were less likely to use respirators in dental procedures (KuKanich, Mulcahy, & Petro, 2025), however Robin et al found that previous experiences of treating zoonotic cases may influence PPE use (Robin et al., 2017). One study found most zoonotic infections occurred in the first three years from graduation (Jackson & Villarroel, 2012),however another found respondents were increasingly likely to report zoonotic infection as their years in practice increased (Lipton et al., 2008). Perceived risk of zoonoses may affect PPE use, where VPs sometimes report being more likely to use PPE if zoonotic exposure is considered likely (Dowd et al., 2013; Robin et al., 2017), however the risk or likelihood is often perceived as low (Robin et al., 2017; Rood & Pate, 2019; Willemsen, Cobbold, Gibson, Wilks, & Reid, 2024) (Anderson & Weese, 2016) (Wright et al., 2008). Positive attitudes toward zoonotic risk reduction were higher among veterinary nurses and those motivated by standard operating procedures (SOPs), positive client perceptions, and personal safety (Robin et al.). Conversely, individuals who viewed time constraints as a barrier to PPE use exhibited lower positive attitude scores (Robin et al., 2017). Some studies found low availability of PPE as a deterrent to use (Dowd et al., 2013; Steele, Mor, & Toribio, 2021), however Robin et al found this had no influence on decisions to use PPE.

The risk profile for zoonotic exposure in companion animal veterinary settings is evolving due to a rising pet ownership, higher volumes of imported rescue animals, and climate-driven shifts in tick-borne disease vectors (Animal and Plant Health Agency, 2022). The UK Health Security Agency reports increasing cases of non-native and travel-related infections in recent years, such as brucellosis and leptospirosis (United Kingdom Health Security Agency, 2025). Though less common, severe zoonoses like Corynebacterium ulcerans-derived diphtheria present ongoing hazards(Animal and Plant Health Agency, 2025b), and contact with companion animals is the most frequently cited exposure for human cases (United Kingdom Health Security Agency, 2025). However, exact transmission rates within veterinary personnel and the proportion contracted through occupational exposure remain unspecified.

Given the inconsistent evidence surrounding PPE compliance and the unique biosecurity challenges of companion animal practice, such as close, prolonged physical contact with pets and high owner interaction, there is a need to understand the professional experiences of these clinicians. This study aimed to explore UK companion animal VPs’ experiences with PPE use in the context of zoonotic risk, identifying key barriers and facilitators to improving PPE practices.

## Materials and Methods

Focus Group Discussions (FGDs) were held at a veterinary referral hospital in the North-West of England facilitated by the primary and secondary researcher to explore experiences of VPs with PPE in the context of zoonoses. FGDs can be used to obtain a broad range of experiences on a topic and help to identify lines of consensus and differences (Green & Thorogood, 2004). They were therefore used to gain a collective insight into the cultural practices of VPs in relation to their experiences with PPE. The topic guide was produced following review of the existing literature and study aims and was screened by a veterinary surgeon and researcher with qualitative experience. This study was approved by the University of Liverpool Institute of Infection, Veterinary and Ecological Sciences Research Ethics Committee in March 2025 (Ref: 16015).

Participants were those working clinically in one referral veterinary hospital. Purposive sampling was used. An email explaining the study and requesting participation was distributed to all veterinarians and nurses at the hospitals via a Professor and Senior Lecturer at the unit and the Head Nurse. Interested participants were sent the participant information sheet and consent form to read and sign before the FGDs.

Four FGDs were held stratified by staff type: one of Veterinary Nurses with current senior management roles; one of Veterinary Nurses without current managerial roles; one of senior Veterinary Surgeons (consultant or resident); and one of junior Veterinary Surgeons (interns). FGDs occurred in person at the hospital in June and July 2025. Number of participants per FGD ranged from three to six. FGDs lasted approximately one hour and were recorded and transcribed using Microsoft Teams. Transcriptions were then checked in Microsoft Word.

Reflexive thematic analysis (Braun & Clarke, 2006) was used inductively to identify key themes to explain VPs’ experience with PPE: one researcher (CC) familiarised themselves with the research, generated initial codes and searched for themes, reviewed, defined and named themes before final write up. The second researcher (JT) reviewed themes. The thematic analysis was used to identify barriers and facilitators to PPE use within this setting.

## Results

Eighteen participants consented to take part in the study (supplementary material). This research identified a central theme with four surrounding themes (Fig. 1), which explain VPs’ experiences with PPE, and have been shaped professionally, socially and culturally. These themes have been used to identify facilitators and barriers to PPE use.

**Figure 1:**
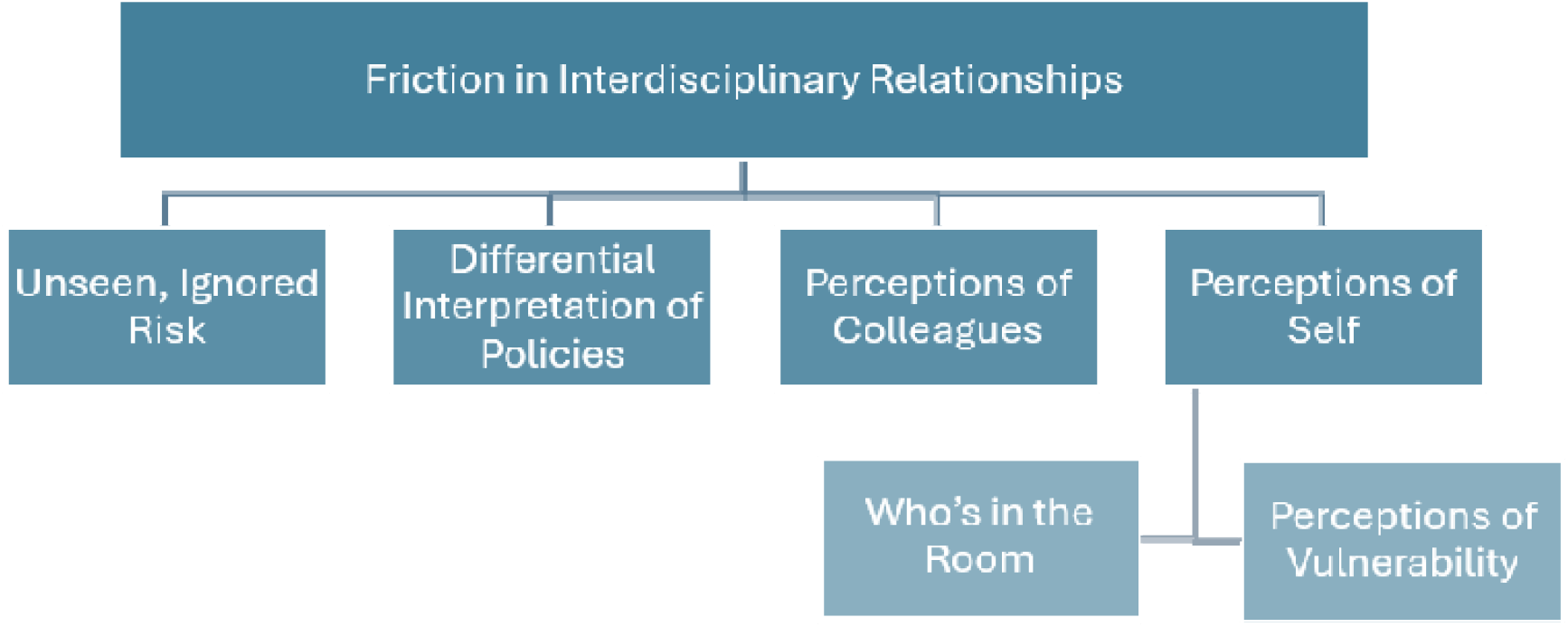
Overview of main themes and sub-themes influencing Personal Protective Equipment (PPE) usage, in relation to zoonoses, among companion animal veterinary staff

### Central Theme: Friction in Interdisciplinary Relationships

The central theme identified is the role of friction in interdisciplinary relationships in shaping VPs’ experiences with PPE. This theme refers to the complex social and professional dynamics between veterinarians and veterinary nurses, where differing views on risk, policy and PPE, contrasting perceptions of each other, and challenges with communication impact PPE decision-making.

Participants identified that PPE use was generally low but agreed that nurses used more PPE than veterinarians. Nurses attributed this to veterinarians’ underappreciation of risk and disregard for protocols based on confidence in the superiority of their own knowledge (Table 1, Quote 1).

**Table 1:** Supportive quotes illustrating identified themes regarding companion animal veterinary staff’s Personal Protective Equipment usage in relation to zoonotic disease risk.

|  |  |
| --- | --- |
| Quote 1 | SVN1: *"their perception of the risk is less because of whatever knowledge or paper at some point that they've read"* |
| Quote 2 | SV3: *"It's probably the nurses (are) much better at using PPE than we are, but also don't know why they're using it, don't understand the disease enough to make a good judgement call which one should have PPE, and which one shouldn't."* |
| Quote 3 | SVN1: *"and as SVN2 said, it is belittling the nurses a lot when they like, "put some gloves on", "oh I don't need gloves"."* |
| Quote 4 | SV3: *"Yeah, people are welcome to wear PPE, but they don't need to put a sign on the door saying that everyone else needs to wear PPE."* |
| Quote 5 | SV1: *"it may also reflect a sort of lack of confidence in transfer of information... You know, in terms of whether they're confident that we would always know if the dog was raw fed or whatever, and whether or not we would tell them"* |
| Quote 6 | SV4: *"We also don't test routinely for antibodies or anything like this, so you would never know if you were exposed"* |
| Quote 7 | SV2: *"I've never used gloves to examine a patient. Unless there is, you know, an open wound or something like that"* |
| Quote 8 | JV2: *"I never would have bothered with gloves even if they came in with like this round belly that you think was full of worms. I just would wash my hands afterwards and never think about it again"* |
| Quote 9 | SVN1: *"I've had lepto (laughs) "*<br>SVN3: *"Have you? Have you actually?"*<br>SVN1: *"Yeah (laughs) "*<br>SVN2: *"That's got ya (laughs)"* |
| Quote 10 | JV6: *"I certainly didn't get any kind of explanation or anything back that I should be tested... but whether or not that could have been maybe highlighted or explained a little bit more, of the risk, afterwards"* |
| Quote 11 | SV3: *"we don't have, for example, a clear policy on imported dogs at the moment that may or may not have brucellosis. So that's very... clinician dependent"* |
| Quote 12 | JVN1: *"we never wear gloves for taking Bloods, general clinical exam... **obviously** we alcohol and clean our hands between each patient"* |
| Quote 13 | SV3: *"it's a bit like sign blindness. If an animal has 10 signs on its door, you're not going to read them all?"* |
| Quote 14 | JVN2: *"But even then, some people won't...see it"*<br>JVN5: *"the vets are the worst for that"* |
| Quote 15 | SV2: "So they are **much** more in contact as well. So I suppose they are also better at using PPE because they see themselves more at risk than **we** are" |
| Quote 16 | SVN1: "Obviously I had to do chest compressions on it and obviously it was just weeing everywhere... And everyone's like "it's got lepto!" and I was like, well, I wasn't going to stop to put PPE (on)! So this is a good example of the fact that I wasn't bothered about me" |
| Quote 17 | JV4: "when you're barrier nursing an inappetent dog when you're trying to tempt it to eat and it just sniffs the latex gloves, it's not going to eat... in some cases (like) that I will just take the gloves off to hand feed it because, It's not going to eat otherwise." |
| Quote 18 | JVN3: "If you cause an issue from patient to patient, you'll get pulled up on it and you'll get in trouble, whereas if you come away with something.... they're not going to tell you off for that. That's kind of like, down to **you** really... they've got no problem pulling you up if it's another patient that it affects" |
| Quote 19 | JVN5: "I'd shit me pants"<br>JVN2: "...I think here they'll be like "when are you coming back?"<br>Participants: (laughing)<br>JVN2: "you work next to a toilet. You'll be fine." |

Veterinary surgeons, both junior and senior, expressed a belief that their knowledge was superior, explaining that nurses “overused” PPE due to poor understanding of disease transmission among their profession (Table 2, Quote 2). This condescension was identified in both nurse FGDs, where participants highlighted their attempts to improve veterinarians’ PPE compliance were often rebuffed (Table 2, Quote 3). Nurses’ attempts to change behaviour further propagated friction, as some veterinarians felt nurses should not be dictating the level of PPE used (Table 2, Quote 4).

**Table 2:** Barriers and Facilitators to Personal Protective Equipment use in companion animal veterinary staff, in 417 the context of zoonoses.

| Barriers | Facilitators |
| --- | --- |
| An absence of initial recognition of risk | Effective inter and intradisciplinary communication |
| A staff culture of patient prioritisation over risk to self | Previous personal experience with serious zoonoses |
| Friction in response to others’ PPE use or lack of |  |
| An overly complex policy, which further divides staff |  |
| Lack of cultural and institutional reaction to occupationally contracted zoonoses |  |

Finally, friction was experienced as a consequence of poor interdisciplinary communication of risk and PPE level from veterinarians to nurses:

> “*SVN1: I suppose the only frustrating thing that we always find is the same, is the lack of communication sometimes between people and that can sometimes be a bit of a cause for concern*”. Veterinarians were aware of this issue and recognised the damage it caused to their interdisciplinary relationship, as well as its subsequent influence on nurses’ PPE decision making (Table 2, Quote 5). Whilst these quotes demonstrate the central theme, friction is also exhibited in the remaining interlinked themes (Figure 1).

### Subtheme: Unseen or Ignored Risk

These are the zoonotic hazards that are either not immediately visible, not fully understood, or consciously overlooked by VPs. Unseen risk arose at times when the presence of a zoonoses was not initially clear, until further diagnostics were performed. In these cases, the absence of obvious clinical signs in animals led to reduced vigilance, inconsistent PPE use, and increased pathogen exposure. When zoonoses were diagnosed, “a quick shift from No PPE to PPE” was described. Nurses and veterinarians described this for both suspected and confirmed zoonotic cases:

> “*SVN1*: “*I think it was just it the same thing that* ***always*** *happens…It doesn’t look like that, you know, like it’s going to be TB [tuberculosis]. They start doing tests and examine them and they’re like, “oh, actually, it could possibly be”. But at that point, the exposures already happened”*.

Risk sometimes went unnoticed as it was intangible or unmeasurable (Table 1, Quote 6). In contrast, where a risk was visible as an immediate threat, PPE use appeared obvious to participants (Table 1, Quote 7).

However, even when a risk was registered by the VP, experiences with PPE did not change, suggesting the risk of infection was being ignored (Table 1, Quote 8). Ignored risk occurred when the hazard was recognised, but its importance is underestimated or downplayed. Reasons alluded to in FGDs were time pressures, discomfort or inconvenience of PPE, perceived low probability of infection, or a workplace culture that normalises exposure. When one participant described contracting Leptospirosis whilst working, they were met with scepticism and bemusement. This propagated a workplace culture that downplays probability of infection and so contributes to ignored risk (Table 1, Quote 9). This effect of institutional culture on risk was seen throughout VPs’ careers. Occupational exposures to known zoonoses were not followed up, normalising the notion such risk can be ignored. This is seen from education to current practice (Table 1, Quote 10).

In contrast, where risk and consequences were considered afterwards, experience with PPE changed. One participant gave an account of a colleague contracting Leptospirosis in general practice. The risk was communicated to other practices in the chain, and the participant reflected on how this experience changed their PPE use:

> “*JV6: we would have meetings every week… it made us aware of this leptospirosis and everybody was full on (laughing) gloves everywhere and I think it just made everyone **think** a bit more*”

Overall, this theme highlights the cognitive and cultural gap between actual risk and perceived risk, where decisions about PPE use were influenced more by social and institutional perceptions of risk and visible, immediate threats, than by invisible hazards like zoonotic infection. Unseen and ignored risk were clear barriers to PPE use, however where risk was openly discussed PPE use increased. Understanding risk is critical for designing interventions that address both knowledge deficits and behavioural norms, shifting PPE use from a reactive, hazard-specific activity to a consistent, preventative practice embedded in routine veterinary work.

### Subtheme: Differential Interpretation of Policy

This refers to the differences in how veterinarians and nurses adhere to policy as well as their overall sentiment towards it, impacting how different VPs experience PPE. The friction between the professions is clear in this theme, where differences lead to clashes.

The Hospital’s self-designed “*traffic light*” protocol states that every patient is assigned a colour and corresponding level of PPE to use, where “*green is just gloves*… amber is gloves and gown, and I think red is like [*the full PPE*]”. Veterinarians viewed this more as a “*guidance*” than a mandate:

> *“JV4: It’s there for everyone to know*…, *but then it’s everyone’s personal decision whether they do so or not”*

Veterinarians viewed IPC policies in general as too numerous, impractical and sometimes illogical. For example, whilst the Hospital’s current policy advises “*full PPE*” for raw fed dogs, which SV3 reports *“not many people do”*, they highlighted there is no clear policy on imported dogs with potential Brucellosis, leading veterinarians to make their own decisions (Table 1, Quote 11).

Nurses however championed the traffic light protocol, with it being referred to as the “*brainchild*” of two of the nurses, stating it is “*there for a reason*”. They often tried to encourage adherence from others, but were frustrated by veterinarians’ response:

> *“JVN5: you’ll put the sign on…clinicians come in, no gloves, swinging the animal round doing neuro exams and you’re like” you’re setting an example to the* ***students***”“.

A point of agreement for both professions was that hand hygiene is often given preference over PPE, despite awareness of protocols (Table 1, Quote 12). Further agreement is in the issue of information overload, with both highlighting that policies require time to read them, as well as the literal information overload when approaching a kennel with multiple signs (Table 1, Quote 13). However, nurses perceived veterinarians as having poorer situational awareness (Table 1, Quote 14).

In summary, differential interpretation of policies shaped VPs’ experience with PPE. Veterinarians viewed policy as an often-excessive guidance, thus acting as a barrier to PPE use. In contrast, nurses championed policies, acting as a facilitator in this cohort. An area of agreement was information overload and prioritisation of hand hygiene over PPE. This common ground could be explored to understand how adoption of PPE in both professions could be improved.

### Subtheme: Perceptions of Colleagues

#### Perceptions of Vulnerability

This subtheme refers to the perception that others are more vulnerable than themselves and was seen in all FGDs. In viewing others, such as those who were immunocompromised, as more vulnerable than themselves, VPs could downplay their own risk and justify not using PPE, linking back to ignored risk in theme one:

> *JVN2: “especially if we consider ourselves to be healthy. Like, I think if I was pregnant or I was immunosuppressed I would think… differently*.”

Nurses were also viewed by both professions as more vulnerable, partly due to increased exposure to patients and bodily fluids, thus explaining their greater PPE use (Table 1, Quote 15). Veterinarians also deemed them more vulnerable due to assumed professional and demographic stereotypes:

> “*JV3: vets…are maybe a little bit more career oriented and not necessarily married with kids all the time. And then the nurses have a different lifestyle (chuckles) and they’re more careful because that part of things, their families, their kids, pregnancies and so on”*

Perceptions of vulnerability were used to explain level of PPE use in themselves and others. The focus on others’ vulnerability ties in with subtheme “Perception of Self”, where one’s self was simply not considered.

#### “Depends who’s in the room”

This subtheme reflects to VPs’ tendency to change behaviour based on who is observing them.

For the nurses, PPE was used partly for “*setting an example for everyone else*”, but also to demonstrate their dedication and professionalism to others:

> “*JVN2: pride comes into it as well*…*you want people to feel you’ve done a good job and you want your patients… It’s like you, your work ethic and I don’t ever want people to say “she’s a rubbish nurse*.”

In contrast, whilst veterinarians also used PPE to set an example, there appeared to be an element of wearing PPE to appease the nurses:

> *“SV3: erm… I honestly think around *Nurse X*, I’m really good. But uh,*
>
> *SV4: yeah around *Nurse X*. Yeah, it’s it’s uh*
>
> *SV3: no our nurses taught me…to be much better”*

Friction between the two professions became obvious, with some disdain at the fact the nurses’ mere presence dictated veterinarians’ behaviour:

> *“SV4: yeah we’re very much on top of it*
>
> *SV3: ruled by our iron leader (laughing)”*

Overall, perceptions of colleagues could be understood as both a barrier and facilitator. Perceiving others as more vulnerable allowed staff to justify omitting PPE for themselves, whereas the need to set an example and show they’ve done “*a good job*” encouraged PPE compliance. Although nurses try to promote PPE use in others, the discontent this built in veterinarians likely negatively impacted their experiences with PPE.

### Subtheme: Perception of Self

This refers to VPs positioning the patient’s needs above their own, both in emergencies and routine care. This stems from a lack of consideration for self, normalisation of occupational exposure, and the perception they are “*invincible*”. These perceptions are reinforced culturally and professionally.

The prioritisation of patients over self was seen in every FGD, demonstrating a commonality between professions (Table 1, Quote 16 and 17). The lack of self-consideration was demonstrated by JVN2:

> *“a patient with E coli, I would wear all the gear, but I would be wearing it to protect my other patients…we kind of forget ourselves, don’t we?”*

This was reinforced professionally, where exposure to bodily fluids, and so risk to self, was normalised:

> *“JVN5: So I get peed on, on a daily basis, so sometimes I’ll just stick one of them (aprons) on and I don’t necessarily need to because I could just get weed on*.*”*

Shared professional values further contributed to lack of self-consideration. When nurses were asked why they see the risk as greater to their patients than themselves, participants referred to essential vocational attributes:

> *“JVN2: Like the nature of people who’ve become nurses, empathy is a massive, massive characteristic-*
>
> *JVN5: Your priority, is your patient, so, your priority is always them. You have to be their advocate”*

The idea that the self is less important was reinforced institutionally, where nurses highlighted the lack of repercussions for contracting a zoonoses themselves compared to spreading a zoonotic nosocomial illness among patients (Table 1, Quote 18). Note the fear of repercussions links to subtheme “Differential Interpretation of Policy” and could explain why nurses followed protocol more tightly. Institutional influence on perception of self was further shown when nurses were asked if they would report contracting a zoonotic disease. They indicated they would not, implying a lack of concern from hospital management (Table 1, Quote 19).

Perceptions of self, which have been shaped socially and institutionally, influenced VPs’ experience with PPE. Lack of self-consideration therefore acted as a barrier. Understanding and challenging these perceptions could provide a useful facilitator.

### Barriers and Facilitators

Collective experiences provided insight into how PPE use in the context of zoonoses has been shaped personally, culturally and socially throughout their career, and how this differed between veterinarians and nurses. Barriers were an absence of initial recognition of risk, a staff culture of patient prioritisation over risk to self, friction in response to others’ PPE use or lack thereof, an overly complex policy, and a lack of cultural and institutional reaction to occupationally contracted zoonoses (Table 2). Facilitators were effective inter and intradisciplinary communication and previous personal experience with serious zoonoses.

## Discussion

Our study demonstrates that interprofessional friction, driven by perceptual differences and poor communication, influenced experiences with use of PPE. Friction in professional relationships was a clear barrier to PPE use, and any interventions are unlikely to be successful unless this is addressed. Interdisciplinary relationships have not been examined in the context of zoonoses previously, however RCVS communication guidelines highlight respectful communication (Royal College of Veterinary Surgeons), and being respected by veterinary surgeons is a factor in staff retention of veterinary nurses (Jeffery & Taylor, 2022). One study found good interprofessional collaboration generally between veterinary nurses and surgeons, but that nurses in university hospitals had poorer experiences than those in general practice, in keeping with our findings (Fontaine, Cadman, Bennet, & McKeegan, 2026).

Previous survey data indicated high perception of risk to self or likelihood of exposure impacted PPE use (Dowd et al., 2013; Robin et al., 2017). However, our study highlights that risk is often unseen, such as in diagnostic uncertainty, therefore risk could not contribute to decisions to wear PPE. Where risk was recognised, it was often actively ignored, which could explain misalignments seen previously between stated perception of risk and lack of appropriate PPE use when managing patients with possible zoonotic illnesses (Wright et al., 2008). The idea that risk is more often registered if it is a tangible, immediate threat was mirrored in research relating to veterinary workplace injuries, where veterinarians expected injuries to involve blood (Furtado, Whiting, Schofield, Jackson, & Tulloch, 2024). In our study nurses identified themselves as more vulnerable due to increased exposure to bodily fluid and so greater users of PPE. Risk may also have been ignored due to placing patient needs above personal safety, a behaviour seen in both veterinary nurses and surgeons. VPs’ under recognition of zoonotic infection risk could be related to a lack of institutional concern or reaction from employers to occupationally contracted zoonoses, which participants had experienced throughout their career. In contrast, one participant reported that their employer shared news of a colleague contracting a zoonotic infection and discussed it in weekly meetings, which prompted a change in their PPE behaviour. This highlights that while personal experience with a zoonotic infection is important, a culture that reinforces risk recognition and response is needed for a wider, cultural shift in PPE use.

While both nurses and surgeons recognised relevant IPC policies, guidelines were broadly viewed as unwieldy and unaligned with workflow demands. The observed discrepancy in compliance, where nurses demonstrated greater protocol adherence than veterinarians, aligns with documented differences in professional role socialisation (Robin et al., 2017). Rather than lack of awareness, veterinarians’ reliance on personal discretion over established guidelines points to a culture of clinical self-reliance (Dowd et al., 2013). This tendency is particularly pronounced when IPC measures are framed as executive management directives rather than clinical necessities, fostering professional pushback (Willemsen et al., 2024). Willemsen et al also recommended an IPC Champion to increase the uptake of IPC policies.

Any policy must consider that overly complex guidance acts as a barrier to PPE use and should be coproduced with those with lived experience with serious zoonoses. Coproduction has been used in the design of IPC policies in human healthcare settings (Shaw et al., 2019). Within One Health approaches to zoonotic disease control, co-production has been hypothesised as a means of integrating scientific evidence with stakeholder experience, improving the relevance, acceptability, and implementation of interventions (Asaaga et al., 2022).

Given the value of risk recognition both individually and culturally, a joint statement released by the HSE, the UKHSA and the RCVS to raise visibility of risk in the profession could support cultural change. With inter and intradisciplinary communication acting as a facilitator and friction between professions a barrier, information regarding PPE instructions for zoonotic cases should be cascaded appropriately to both nurses and veterinarians.

Based on the study findings we have the following recommendations regarding PPE use in veterinary hospitals. Firstly, organisations must move away from rigid, administrative mandates by actively collaborating with veterinary staff to co-produce simplified, user-friendly IPC and PPE guidelines. Involving peer advocates ensures protocols remain grounded in practical workflow demands. Redesigned guidance should be piloted within small cohorts prior to hospital-wide implementation to identify practical barriers, gather feedback, and refine protocols for maximum acceptability. Second, addressing veterinary resistance requires framing these updated measures explicitly as direct clinical necessities rather than executive management directives. Targeted educational initiatives should specifically address veterinarian autonomy, bridging the gap between independent clinical judgment and standardised safety protocols to counter the tendency to bypass established rules. Finally, fostering long-term behavioural change depends on establishing a risk-aware culture supported by high-level endorsement and equitable communication. A joint statement released by regulatory and public health bodies (specifically the HSE, UKHSA, and RCVS) would elevate the visibility of zoonotic risks across the entire profession. At the practice level, this must be complemented by a transparent, interdisciplinary communication strategy that cascades PPE instructions simultaneously and appropriately to both veterinary nurses and surgeons, eliminating information silos and minimising interprofessional friction.

A strength of this study is that it is the first qualitative study investigating VPs’ experiences with PPE in the context of zoonoses in the United Kingdom. Whilst quantitative research identifies prevalence and some barriers and facilitators (Robin et al., 2017), qualitative methods enable a deeper exploration of how and why these factors shape everyday practice. This approach provides contextual insights into participants’ experiences and decision-making that survey data alone cannot capture (Green & Thorohood, 2004), allowing for a more comprehensive understanding of PPE use in zoonotic risk management. Our study has limitations. Participants that volunteered were likely to be more interested in PPE and IPC, introducing selection bias. Social desirability bias may also have affected results, as research in IPC practices in human healthcare professionals has shown a discord between self-reported and actual behaviour (Jenner et al., 2006). We mitigated this by stratifying FGDs by profession and seniority. Finally, all participants were recruited from a single tertiary hospital. Participants had had prior general practice experience which may shape their behaviours, however results cannot be generalized to general practice or other clinical settings.

This study provides insight into VPs’ experiences with PPE in the context of zoonoses. By examining both collective and individual perspectives, the study highlights facilitators, such as communication and personal experience, and barriers, such as patient prioritisation, ignorance of risk, overly long policies and interdisciplinary friction, to PPE usage. Together, these insights inform practical recommendations to increase PPE use.

## Data Availability

All data produced in the present study are available upon reasonable request to the authors

## Acknowledgements

We would like to thank the staff of the anonymous referral hospital used in this study taking the time out of their busy schedules to participate in this research.

## Conflict of Interest Statement

There are no conflicts of interest to be declared.

## Notes

### Competing Interest Statement

The authors have declared no competing interest.

### Author Declarations

This study was approved by the University of Liverpool Institute of Infection, Veterinary and Ecological Sciences Research Ethics Committee in March 2025 (Ref: 16015).

