## Supplementary Material for "Understanding Personal Protective Equipment Use Among Companion Animal Veterinary Staff in the Context of Zoonoses: A Qualitative Study"

**Supplementary Materials**

| ***Focus Group*** | ***Pseudonym*** | ***Occupation*** |
| --- | --- | --- |
| ***Focus Group 1*** | *SVN1* | *Veterinary Nurse with Senior Managerial Experience* |
|  | *SVN2* | *Veterinary Nurse with Senior Managerial Experience* |
|  | *SVN3* | *Veterinary Nurse with Senior Managerial Experience* |
| ***Focus Group 2*** | *JVN1* | *Veterinary Nurse* |
|  | *JVN2* | *Veterinary Nurse* |
|  | *JVN3* | *Veterinary Nurse* |
|  | *JVN4* | *Veterinary Nurse* |
|  | *JVN5* | *Veterinary Nurse* |
| ***Focus Group 3*** | *SV1* | *Resident Veterinary Surgeon* |
|  | *SV2* | *Consultant Veterinary Surgeon* |
|  | *SV3* | *Consultant Veterinary Surgeon* |
|  | *SV4* | *Consultant Veterinary Surgeon* |
| ***Focus Group 4*** | *JV1* | *Intern Veterinary Surgeon* |
|  | *JV2* | *Intern Veterinary Surgeon* |
|  | *JV3* | *Intern Veterinary Surgeon* |
|  | *JV4* | *Intern Veterinary Surgeon* |
|  | *JV5* | *Intern Veterinary Surgeon* |
|  | *JV6* | *Intern Veterinary Surgeon* |

Supplementary Table 1: Focus Group Participant Information
